# CLINES: Clinical LLM-based Information Extraction and Structuring Agent

**DOI:** 10.64898/2025.12.01.25341355

**Authors:** Zongxin Yang, Hongyi Yuan, Raheel Sayeed, Amelia Li Min Tan, Enci Cai, Mohammed Moro, Xiudi Li, Huaiyuan Ying, Nicholas Brown, Griffin Weber, Sheng Yu, Isaac Kohane, Tianxi Cai

## Abstract

**Objective:** Clinical narratives in electronic health records (EHRs) hold diagnostic, therapeutic, and temporal information often missing from structured fields; manual chart review remains the de facto standard but is slow, costly, and variable. We aimed to develop and evaluate CLINES, a pipeline that extracts and normalizes clinical concepts from free-text notes into ontology-grounded, auditable, schema-ready data without task-specific fine-tuning.

**Methods and Analysis:** We developed CLINES, a modular multi-step pipeline: semantic chunking of long notes; extraction by reasoning-capable large language models; attribute assignment (assertion, value+unit); UMLS normalization; explicit and relative date resolution; aggregation into an i2b2-style schema. Evaluation was conducted without task-specific supervised training on de-identified MIMIC-III, 4CE, and CORAL (breast, pancreas) notes. Comparators: rule/lexicon systems, transformer encoders, single-prompt LLM baselines, alongside chain-of-thought LLMs and full-size clinical NER encoders (BERT-base, GatorTron-base). Outcomes were F1 for entity, assertion, value+unit, and date processing, reported with 95% bootstrap CIs and paired-bootstrap FDR-corrected tests; inter-annotator agreement, hallucination taxonomy, cost, and component ablations are in Supplementary Materials.

**Results:** Across all datasets and tasks, CLINES ranked first on every task-dataset combination. F1 ranges were 0.69–0.87 (entity), 0.84–0.93 (assertion), 0.77–0.90 (value+unit), and 0.73–0.79 (date, where gold annotations were available); per-dataset values are in Supplementary Materials. Gains over the strongest single-prompt LLM were +0.21–0.38; transformer encoders trailed by +0.28–0.68 F1 on entity extraction. Performance remained stable as notes lengthened, while transformer baselines lost recall. Code-field gains and most assertion/value comparisons were significant at *p*_FDR_ *<* 0.05 on paired-bootstrap tests; mention-level inter-annotator F1 on a cross-annotated subset was 0.82 (assertion *κ* = 0.85); a seven-class taxonomy on 700 stratified false positives found 54% of apparent false positives to be annotation gaps rather than true hallucinations.

**Conclusion:** CLINES translates narrative EHR text into ontology-grounded, auditable, schema-ready data, offering a practical route to scale chart-review-like extraction for cohort discovery and real-world evidence. CLINES substantially reduces, but does not eliminate, the need for manual review: the residual hallucination quantified here means human verification remains necessary for safety-critical use. CLINES is model agnostic–open and closed models can be substituted to meet cost, performance, and privacy goals.

**Summary box:** *What is already known on this topic:* Free-text clinical notes hold diagnostic, temporal, and quantitative detail absent from structured EHR fields; rule-based systems and fine-tuned transformer encoders detect entities but normalize and attribute them inconsistently, and single-prompt large language models hallucinate and miss instructions on long notes.

*What this study adds:* CLINES, a training-free multi-step large-language-model pipeline, produces UMLS-normalized, attributed, time-stamped output that substantially outperforms single-prompt and chain-of-thought LLMs (+0.37 to +0.54 code F1) and full-size clinical encoders; the gains hold across open- and closed-weight backbones and are quantified with confidence intervals, inter-annotator agreement, a hallucination taxonomy, per-note cost, and component ablations.

*How this study might affect research, practice, or policy:* CLINES offers an auditable, model-agnostic route to scale chart-review-like extraction for cohort discovery and real-world evidence, while the quantified residual hallucination rate makes explicit that human verification remains necessary before safety-critical use.

## 1. Introduction

Clinical narratives in electronic health records (EHRs) are indispensable for understanding patient status, informing decision support, and enabling biomedical research. Narrative notes—spanning discharge summaries, progress notes, imaging and operative reports—capture observations, differential diagnoses, treatments, test results, and clinician reasoning that structured fields rarely encode. Expressions such as “intermittent chest pain worsening with exertion but relieved by rest” or “likely drug-induced hepatitis, possibly from isoniazid” capture nuance that structured fields miss; measurements like cardiac ejection fraction values or tumor size often do not fit neatly into predefined problem lists and billing codes yet are central to clinical interpretation^1,2^. Interpretation is not just an important task of clinical interpretation for immediate clinician decisions. It also is the translation of this text to codified terminologies (e.g., CPT codes) or numerical representations (e.g., lab result tables) to drive tasks in epidemiology, automated decision support, reimbursement, and trial eligibility, among other downstream uses of electronic healthcare data.

Not only are the structured EHR entries incomplete or inaccurate, but conventional natural language processing methods—such as standard named entity recognition—often fail to capture the nuance of clinical reasoning, contextual qualifiers, or temporal relationships in notes^3^. Even when structured codes or extracted entities are available, they may not reflect whether a condition was truly present, a medication was taken, or a disease state had recurred^4^. Consequently, high-quality labels in practice still rely on chart review: investigators read notes to adjudicate diagnoses and verify supporting evidence (medications, symptoms, tests). Yet, chart review is slow, costly, and variable across annotators; double abstraction is frequently required to ensure quality. This persistent reliance highlights the need for automated, scalable approaches that can more effectively extract, structure, and standardize clinical information consistently while leaving an auditable trace.

Classical approaches to clinical natural language processing have attempted to automate extraction and structuring of information from narrative notes but face important limitations. Rule- and lexicon-driven systems, such as cTAKES^5^, MetaMap^6^, and MedLEE^7^, map entities to controlled vocabularies (e.g., the Unified Medical Language System), using handcrafted rules and dictionaries for concept recognition, negation detection, and temporal normalization. While these pipelines established the foundation for clinical NLP, they struggle with shorthand, spelling variants and errors, as well as non-standard grammar common in clinical notes, and they require substantial manual engineering to adapt across institutions and use cases^1,8^. Between these rule-based systems and modern large language models, a generation of supervised neural methods advanced clinical entity recognition and normalization—BiLSTM-CRF sequence taggers and, subsequently, domain-pretrained transformer encoders such as BioBERT and ClinicalBERT^9–11^. These improved span detection but require sizeable task-specific labeled corpora, remain brittle to phrasing variation and institutional shift, and typically address entity detection in isolation rather than joint normalization, attribute, and temporal extraction.

Building on these limitations of rule- and lexicon-driven systems, transformer-based large language models—such as GPT-4^12^ and Med-PaLM^13^—have reshaped NLP and show promise for clinical tasks including entity and relation extraction, summarization, and question answering^14^. Domain-scaled models, such as GatorTron^15^ and Clinical-MobileBERT^16,17^, report strong performance on bench-marks such as i2b2 concept extraction^18^, and instruction-tuned assistants can generalize in zero- and few-shot regimes^19^. Yet, important barriers remain for safe clinical deployment extraction: ontology grounding to UMLS/SNOMED CT is often imprecise with occasional hallucinated codes^20,21^; context and output-length limits complicate reasoning across long clinical notes^22,23^; and in real-world datasets (e.g., from the 4CE consortium), note lengths frequently exceed such limits, hindering cross-sectional reasoning^24^. As a result, single-pass prompting remains unreliable for chart-review-like decisions at scale.

To address these gaps, we present CLINES—a Clinical LLM-based Information Extraction and Structuring agent that brings the consistency of chart review into an automated pipeline. Unlike prior systems limited by handcrafted rules or naive single-pass prompting, CLINES integrates several innovations: informed segmentation for processing long notes; reasoning-capable LLM extraction of entities and relations; attribution of clinical context (e.g., negation, experiencer, dosage, laboratory values); normalization to existing ontologies (e.g., UMLS); temporal alignment of events; and reconciliation into an i2b2-style schema to facilitate downstream integration. By uniting these components, CLINES represents a substantial step forward in clinical information extraction, with improvements over both classical NLP pipelines and baseline LLM prompting. Its improvements are validated using real-world EHR notes spanning multiple disease areas and institutions, underscoring accuracy across the disease areas and institutions evaluated; we nonetheless temper claims of broad generalizability, as several common note types (e.g., discharge summaries, operative and radiology reports) remain unevaluated. Equally important, CLINES is engineered to enable different generative models to be substituted within the workflow as dictated by user or institutional preferences regarding cost, property, performance and security. CLINES assumes de-identified, plain-text notes as input; upstream de-identification, format normalization, and document segmentation lie outside the present scope and are addressed as deployment considerations in the Discussion.

## 2. Methods

### 2.1. CLINES architecture and modules

To leverage the superior performance of large language models within manageable context length, CLINES is designed as a four-step pipeline. Instead of an end-to-end approach–where LLMs often hallucinate or miss instructions in lengthy or complex notes–each key task is modularized and paired with carefully designed prompting strategies. The workflow is illustrated in Figure 1.

**Figure 1:**
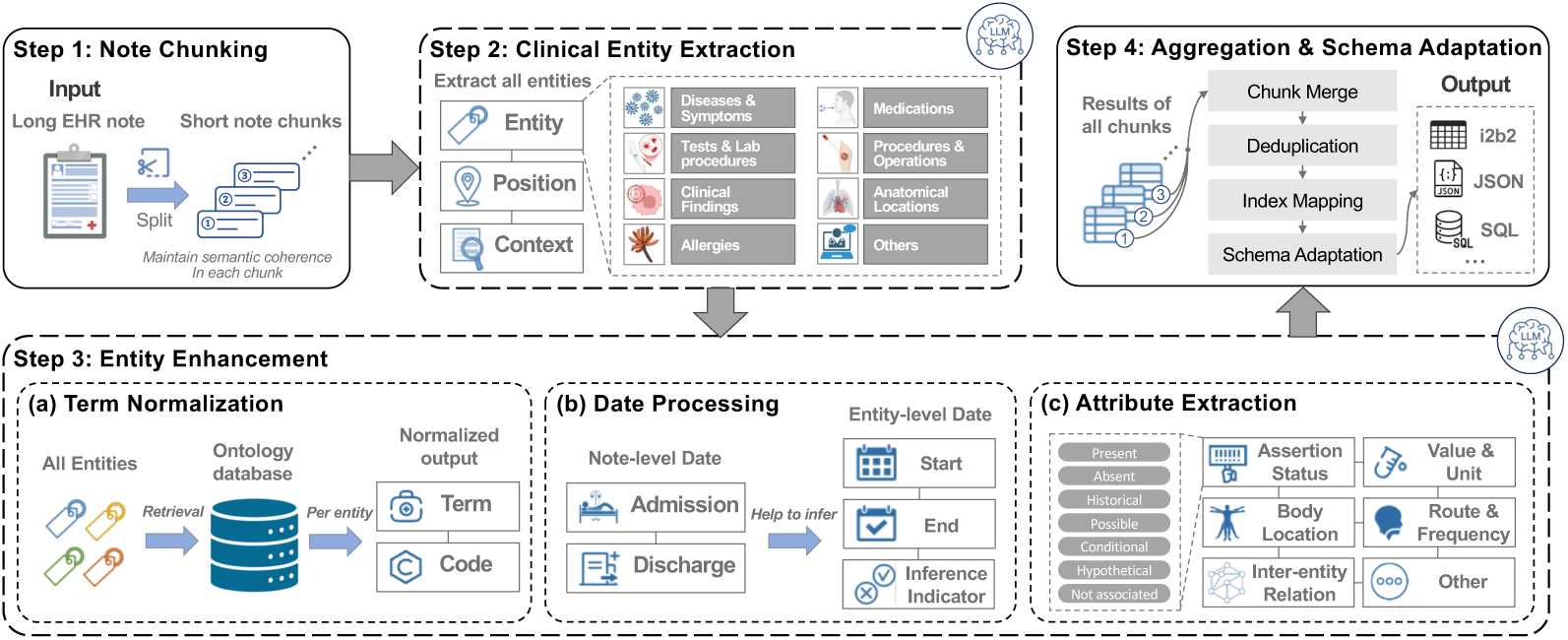
Overview of CLINES. Step 1, segments long clinical notes into semantically coherent chunks to constrain LLM’s context window and improve robustness. Step 2, from each chunk, extract clinical entities with broad coverage, including medications, diseases, and laboratory tests. Step 3, for each entity, uses local context to: (a) normalize to an ontology term with a unique code; (b) for event-type entities, extract or infer start and end dates, when imprecise; and (c) infer assertion status and capture clinically relevant modifiers, e.g., for medications: dose, unit, frequency, and route; for tests: numeric values (with units); for diseases: activity status and other pertinent attributes. Step 4, reconciles entity-level information across all chunks and exports results in user-specified formats (e.g., i2b2, JSON, SQL).

#### Step 1 – Note chunking

Input notes are tokenized with TikToken^25^ and segmented using SemChunk^26^ into semantically coherent fragments of roughly 768 tokens, preserving clinical meaning while fitting LLM context windows. Chunks are processed in parallel to increase throughput. Core metadata (e.g., admission or discharge dates) is retained in all chunks to support temporal alignment. Chunks are non-overlapping; the chunking and overlap policy are detailed in Supplementary Materials.

#### Step 2 – Clinical entity extraction

Each chunk is processed to identify relevant mentions (e.g., symptoms, findings, medications) in narrative order, with positions and contexts preserved. Maintaining order captures clinician reasoning flow while isolating core concepts reduces redundancy and simplifies downstream relation, attribute, and temporal extraction.

#### Step 3 – Concept and attribute enrichment

**(a) Term normalization:** Extracted mentions are mapped to ontology concepts (e.g., UMLS) using LLM-assisted disambiguation and SapBERT-based retrieval, where SapBERT^27^ provides biomedical embeddings; candidates are ranked by cosine similarity. **(b) Date processing:** Explicit dates are normalized to YYYY-MM-DD. Relative or implicit references (e.g., “three days post-admission”) are inferred using anchors such as admission and discharge timestamps; ambiguous dates are flagged as inferred. **(c) Attribute extraction:** For each normalized concept, the pipeline captures assertion status (seven classes: Present, Absent, Historical, Possible, Conditional, Hypothetical, Notassociated; see Supplementary ‘JSON Schema’), numerical values with inferred units, medication route and frequency, anatomical location, and relations between entities (e.g., infection “has symptom” presyncope, “managed by” rehydration). This enriched representation supports downstream phenotyping, predictive modeling, and real-world evidence generation^28,29^. Relation extraction is deployed but not benchmarked here; gold relation annotations are unavailable in all four corpora.

#### Step 4 – Aggregation and schema adaptation

Outputs are reconciled across chunks by re-indexing chunk-local entity tags into a document-global namespace and joining per-step attribute and date outputs back to entity records by global tag; relative-date expressions are resolved against the document’s admission timestamp. Harmonized outputs are mapped to a standardized schema (e.g., i2b2) or a target schema specified by the use-case, then exported to CSV, JSON, or SQLite for downstream analysis. Module-level dataflow is summarized in Figure 2; the reconciliation algorithm is detailed in Supplementary Materials.

**Figure 2:**
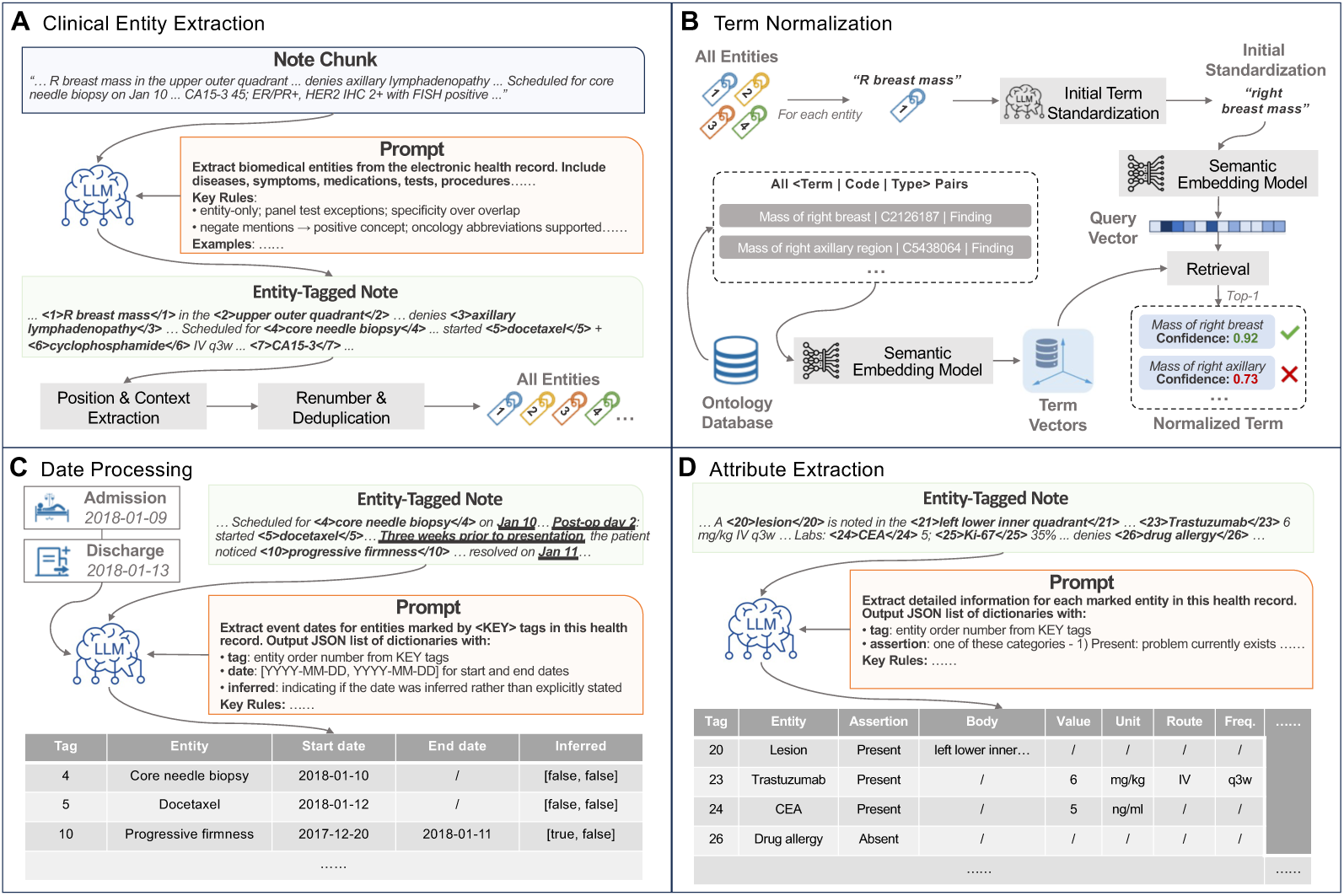
CLINES LLM-based module workflow. (A) Within each note chunk, the LLM tags clinical entities; post-processing yields an entity list with source offsets and a surrounding context window. (B) The LLM performs preliminary normalization, after which an embedding-based retriever queries the ontology database to return the nearest term, concept code, and semantic type. (C) Explicit dates/times are extracted from local context; when documentation is vague, admission/discharge metadata are used to resolve relative or fuzzy expressions to the most plausible calendar dates. (D) Context is used to capture modifiers (e.g., for medications: dose, unit, frequency, route; for tests: numeric values and units; for diseases: activity status), and to infer latent modifiers implied by panel tests (e.g., default units). All machine-inferred fields are flagged as “inferred” to facilitate audit and downstream use.

### 2.2. Implementation and LLM configuration

CLINES is model-agnostic; we employed three instruction-finetuned frontier LLMs with identical decoding settings: Llama-3.1-405B (temperature 0.6, max output 8192 tokens; locally hosted on 8 × H100 GPUs, FP8 precision via SGLang), GPT-4o (temperature 0.6, max output 8192), and o3-mini (medium reasoning effort, max output 32768). Notes were chunked to ≤768 tokens of raw text per request. With GPT-4o, mean prompt tokens were ≈ 8088.25 and mean completion tokens ≈ 2795.25 per chunk; with o3-mini, mean completion tokens increased to ≈ 24334.50. GPT-4o and o3-mini were accessed via API (no local GPUs). Hosted-API calls (GPT-4o, o3-mini) ran on HMS Information Technology’s managed Azure AI service (HMS Azure AI) on de-identified clinical text, with no data retained or used to train provider models; the open-weight backbone was served on-premises. Per-step invocation, token, and cost detail appear in Supplementary Materials.

CLINES outputs were exported to the i2b2 schema, and a read-only web demonstrator populated with results from the credentialed-access CORAL dataset was deployed; access is described in the Data and Code availability section.

### 2.3. Study design and data

All corpora were de-identified and accessed under the respective data-use agreements and governance requirements of each dataset; no patient contact, intervention, or access to identifiable private information occurred. Annotator credentialing, IRB status, and Common-Rule applicability are documented in the Ethics Approval section.

#### MIMIC-III^30^

from 2,065,096 critical-care notes, 964 notes were sampled and segmented into 2,000 paragraphs. Twenty-four paragraphs were manually annotated (one expert per paragraph), yielding 448 clinical phrases: 235 body-location mentions, 128 modifiers (e.g., assertion status), and 144 numerical values with units. Acronym–expansion pairs were treated as single phrases; narrative “purpose” attributes were excluded from quantitative analysis.

#### 4CE24

de-identified clinical notes from patients with obesity and a history of COVID-19, obtained via the Harvard Medical School DBMI Data Portal (see Data Sharing). Of 56 notes, 21 were annotated, producing 5,731 entity mentions with temporal information and attributes; mean document length 1,466 words (SD 636).

#### CORAL^2^

56 de-identified oncology reports (breast or pancreatic cancer). Twenty-nine notes were annotated and partitioned into two subsets: CORAL–Breast (13 documents, 3,900 entity records; mean 1,997 words) and CORAL–Pancreas (16 documents, 4,717 entity records; mean 1,906 words). Across the annotated 4CE + CORAL subsets (14,348 entity records), 2,517 (17.5%) contained both a numerical value and a unit, and 5,680 (39.6%) carried explicit temporal information.

### 2.4. Annotation protocol

Five annotators followed a consensus protocol balancing accuracy and efficiency: (i) extract core entity spans while excluding modifiers to avoid redundancy; (ii) infer temporality and assertion status from surrounding context (e.g., “diagnosed with lung cancer 5 months ago,” “rule out pneumonia”); (iii) expand abbreviations and non-standard terms to full medical concepts, later normalized via SapBERT^27^; (iv) infer missing units for numerical values. Annotation was supported by a custom React-based tool enabling interactive review, conflict resolution, and structured export. All annotators received unified training with shared examples; an independent annotator audited a sample for accuracy and consistency. Throughout, *words* denotes whitespace- and punctuation-delimited tokens (stopwords included; a numeric value together with its unit is counted as a single word), and a *clinical phrase* denotes a contiguous span corresponding to a single UMLS-mappable concept or a single value-plus-unit, as marked by annotators.

### 2.5. Inter-annotator agreement

To quantify annotator variability, we performed a structured cross-annotation study on three notes per dataset (4CE *n* = 3; CORAL *n* = 3; total 2,675 candidate-mention rows): a second annotator, blinded to the original assignment, independently re-annotated each AI-suggested draft. We report mention-level agreement as set-based F1, assertion agreement as Cohen’s *κ* on a 5-class collapsed label space, and value, unit, and date agreement as exact-match rates with denominators defined per field (full agreement-metric definitions, pre-specified structural-disambiguation patterns, and per-note breakdown in Supplementary Materials). For MIMIC-III, where each paragraph was annotated by a single expert, F1 is reframed as agreement-with-annotator.

### 2.6. Baselines

To contextualize performance, we compared classical rule-/lexicon-driven pipelines and compact Transformer encoders, representing the two dominant traditions in clinical NLP.

#### Rule-/lexicon-based systems

cTAKES^5^ implements sectioning, sentence boundary detection, tokenization, POS tagging, morphological normalization, UMLS mapping, and ConText for negation, uncertainty, and experiencer; shallow parsing and dependency analysis link entities to attributes. MetaMap^6^ performs POS tagging, variant generation, composite noun-phrase formation, and UMLS matching with heuristic scoring plus statistical word-sense disambiguation; it does not natively extract numerical values, units, or temporal relations.

#### Transformer encoders

Clinical-MobileBERT and Clinical-DistilBERT^16,17^ fine-tuned on i2b2-2010 concepts^18^ were included as parameter-efficient clinical NER baselines; in addition, two larger off-the-shelf clinical NER encoders were evaluated: BERT-base clinical NER (110M parameters) and GatorTron-base (345M parameters); model identifiers are listed in Supplementary Materials; evaluations were limited to entity extraction because they do not natively support ontology normalization, attribute extraction, or temporal reasoning. Model configurations and hyperparameters followed the authors’ implementations.

#### Single-prompt LLMs

Llama-3.1-405B and GPT-4o were also evaluated in a single-call setting using a compact prompt specifying the target schema and strict JSON output; notes were chunked only to satisfy context limits, and each chunk was processed once without retrieval, tools, cross-chunk aggregation, or multi-step reasoning.

### 2.7. Evaluation metrics and protocol

In the training-free setting (prompt engineering with few-shot in-context examples, retrieval-augmented normalization, and post-processing rules; not zero-shot in the strict sense), we evaluated: entity detection (phrase-level precision, recall, F1 with exact-span match); attribute extraction (joint exact match of span and attribute for body location, assertion status, value+unit); date extraction and normalization (agreement with gold ISO dates; relative dates correct if the resolved absolute date matches the gold anchor). For Clinical-MobileBERT/Clinical-DistilBERT we report only NER; for cTAKES and MetaMap we report concept identification and assertion where available. End-to-end structuring, normalization, attribute extraction, and date handling are compared only across systems that produce those outputs. Completed MI-CLAIM and TRIPOD+AI reporting checklists are provided in the supplementary materials.

### 2.8. Patient and Public Involvement

This study used previously collected, fully de-identified electronic health record datasets and did not involve prospective recruitment or direct contact with patients. Patients and members of the public were therefore not involved in setting the research question, in the design, conduct, or interpretation of the study, or in the preparation of this manuscript. As the study analyses only de-identified secondary data, individual patients could not be identified and dissemination of results to study participants is not applicable. Future translational deployment of CLINES in care settings would benefit from patient and public involvement to inform acceptable use, transparency, and oversight, as noted in the Discussion.

## 3. Results

### 3.1. Benchmark overview

Across four corpora (MIMIC-III, 4CE, CORAL–Breast, CORAL–Pancreas) and four extraction tasks (entity, assertion status, value+unit as a single field, and date processing with start–end joint correctness), CLINES is benchmarked against rule/lexicon pipelines, transformer encoders, and single-prompt LLMs (Figure 3A–D). CLINES (o3-mini) ranks first on every available task–dataset combination. Relative to the strongest single-prompt LLM baseline in the original benchmark cohort (GPT-4o single-prompt), absolute F1 gains are +0.21–0.33 for entity extraction, +0.21–0.28 for assertion status, +0.27–0.38 for value+unit, and +0.26–0.33 for date processing. Versus transformer encoders (entity only), CLINES improves F1 by +0.28–0.68 across datasets. Non-LLM baselines trail on entities and do not produce value+unit or date outputs; among them, only cTAKES yields assertion labels.

**Figure 3:**
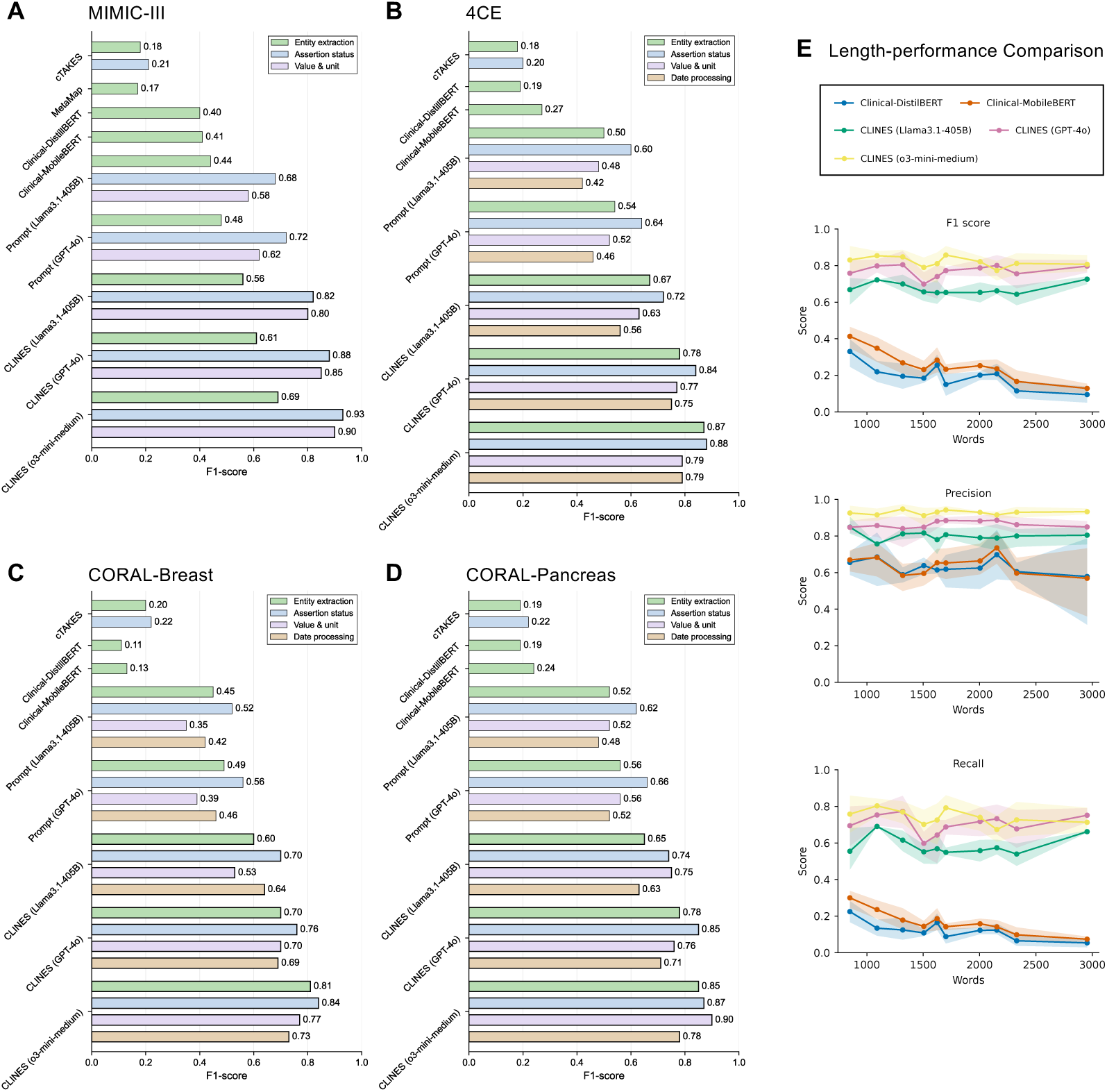
Performance of CLINES across datasets and tasks. (A–D) Results on MIMIC-III, 4CE, and CORAL—Breast/Pancreas comparing CLINES with traditional rule-based NLP, lightweight transformer-based baselines, and LLMs with single-step prompt engineering across six extraction tasks (treating value+unit as a single field and evaluating start+end date as joint correctness). The metric is F1 score. (E) CLINES versus transformer-based baselines as note length/complexity increases, reported as F1, precision, and recall. Notes are grouped into quantile-based length bins; markers indicate the bin-median word count; lines show bin-mean scores; shaded ribbons denote 95% bootstrap confidence intervals.

All primary F1 comparisons additionally carry 95% percentile bootstrap confidence intervals (*B* = 2000 document-level resamples) and paired-bootstrap, FDR-corrected pairwise tests; the per-field point estimates, full bootstrap CIs, and pairwise test results are tabulated in the Supplementary Materials, where Code-field gains and most assertion/value cells are significant at *p*_FDR_ *<* 0.05 and date-column differences are not reliably resolvable at the available sample sizes.

### 3.2. Across corpora and variants

The three CLINES variants maintain a consistent ordering—o3-mini *>* GPT-4o *>* Llama-3.1-405B—for entities, assertions, and value+unit, with all three delivering date extraction on 4CE and CORAL. Single-prompt LLMs outperform rule/lexicon and encoder baselines yet remain below CLINES on every task. Quantitatively, hosted GPT-4o trails o3-mini-medium by approximately 10 pp code F1 and the open-weight Llama-3.1-405B trails by approximately 20 pp.

### 3.3. Note-length analysis

Across quantile-based length bins (∼1–3k words), all CLINES variants show stable F1, precision, and recall (Figure 3E). Transformer-encoder baselines decrease with note length and complexity, driven primarily by falling recall, which remains substantially lower than CLINES across bins.

### 3.4. Expanded baselines, cost, and ablation

Throughout the analyses below we adopt CLINES (o3-mini-medium) as the reference configuration and report the per-field detail on the three consensus-annotated corpora (4CE, CORAL–Breast, CORAL–Pancreas); MIMIC-III is discussed separately because of its single-annotator-per-paragraph protocol. On a cross-annotated subset (three notes per dataset; 2,675 candidate-mention rows), pooled mention F1 was 0.82 and assertion Cohen’s *κ* was 0.85; a post-stratified analysis of false-positive predictions estimated that 54.3% were annotation gaps rather than true model errors, with no individual true-hallucination category exceeding 17% — full inter-annotator-agreement and hallucination-taxonomy detail are in Supplementary Materials. (i) Per-note cost (Figure 4B): $0.21 (o3-mini-medium hosted) versus $0.97 (o3-mini single-prompt) versus ≈ 0.4 GPU-hours (Llama-3.1-405B FP8, 8×H100). (ii) Component ablation (Figure 4C): SapBERT-based UMLS normalization was load-bearing (≈ 0.43 code-F1 loss when removed); cross-chunk reconciliation, semantic chunking, and the date module contributed smaller gains. (iii) Architecture comparison against alternative prompting strategies and off-the-shelf clinical encoders (Figure 4D): the architecture-attributable gain over o3-mini single-prompt and GPT-4o chain-of-thought was +0.37 to +0.54 code F1; full-size clinical encoders (BERT-base 110M; GatorTron-base 345M) matched mention detection (F1 0.84–0.89) but emit no normalized codes, assertion, value+unit, or dates.

**Figure 4:**
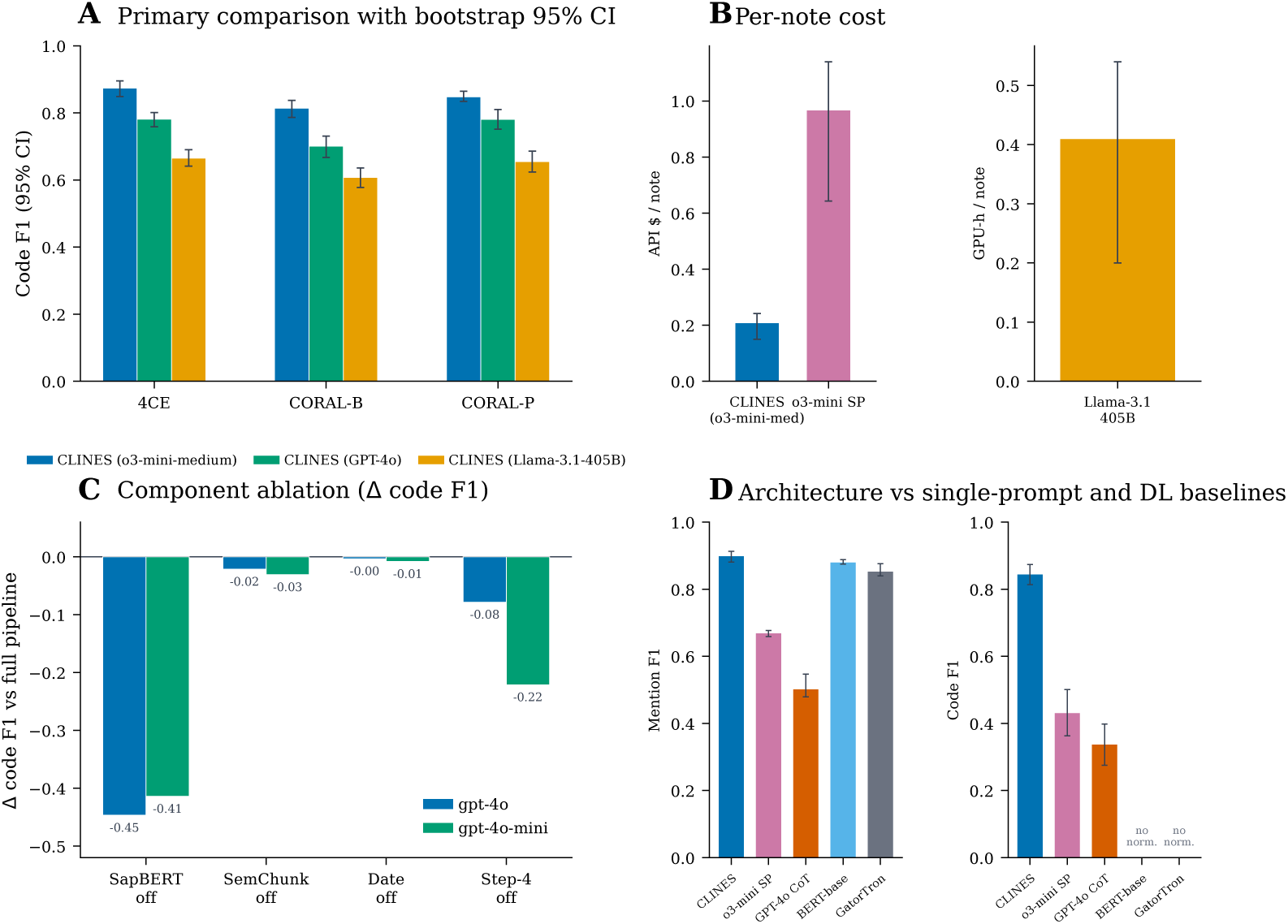
Robustness, cost, and component contributions. (A) Primary CLINES (o3-mini-medium) code-F1 comparison against backbone alternatives (CLINES (GPT-4o), CLINES (Llama-3.1-405B)) with 95% percentile bootstrap confidence intervals. (B) Per-note cost: hosted-API dollars per note (CLINES (o3-mini-medium) versus the same backbone in single-prompt mode) and local GPU-hours per note (Llama-3.1-405B FP8 on 8×H100); the dollar and GPU-hour axes are not directly comparable. (C) Component ablation (change in code F1 relative to the full pipeline, mean of three datasets) for two backbones (GPT-4o and GPT-4o-mini); removing SapBERT-based UMLS normalization dominates on both backbones. (D) CLINES versus single-prompt LLM baselines (o3-mini single-prompt, GPT-4o chain-of-thought) and full-size clinical deep-learning encoders (BERT-base clinical NER, GatorTron-base); the encoders emit entity spans only and produce no UMLS codes, so their code F1 is undefined.

## 4. Discussion

### 4.1. Principal findings

The CLINES pipeline advances the automation of structuring clinical narratives, transforming narrative notes into standardized data formats that can be readily ingested into relational databases or analytical data warehouses. By design, CLINES bridges the longstanding divide between narrative clinical documentation and structured queryable formats, enabling scalable solutions for cohort discovery, real-world evidence generation, and clinical decision support. Critically, its robustness across diverse note types and lengths positions CLINES to unlock the vast reservoirs of unstructured text in EHRs. At scale, this capability can accelerate translational research, enhance population health monitoring, and power next-generation learning health systems by making rich narrative data systematically analyzable.

### 4.2. Backbone model considerations

Our findings reaffirm that backbone choice materially affects downstream performance. CLINES powered by o3-mini/GPT-4o consistently outperformed CLINES powered by Llama-3.1-405B, especially for contextual inference (e.g., implicit unit identification). This should not be interpreted as a direct head-to-head comparison — o3-mini/GPT-4o are proprietary models with broad pretraining on diverse text, whereas Llama-3.1-405B is open-source with more limited access to real-world clinical text. Performance disparities are therefore expected, underscoring the trade-off between accuracy and accessibility. Quantifying this trade-off: within CLINES, hosted GPT-4o trails reasoning-tuned o3-mini-medium by approximately 10 pp code F1 and the open-weight Llama-3.1-405B trails by approximately 20 pp; nevertheless, even the weakest CLINES backbone (Llama-3.1-405B) leads the strongest non-CLINES single-prompt baseline by ≈ 0.15–0.25 code F1, so the architecture’s contribution dominates the backbone’s, and institutions can make a transparent accuracy/cost/privacy trade-off. A key advantage of CLINES is flexibility: the pipeline can be adapted to most LLMs so users can choose the backbone that aligns with performance, cost, and governance priorities. With the recent availability of GPT-OSS, CLINES can be straightforwardly updated to leverage such advances while maintaining accessibility.

### 4.3. Positioning versus existing approaches

Traditional clinical NLP—predominantly rule- or dictionary-driven—faces limited portability and high maintenance across institutions and note types; systems such as cTAKES and MetaMap emphasize lexicon/UMLS mapping and context heuristics but provide partial coverage for attributes or temporal relations. Compact transformer encoders fine-tuned for clinical NER are efficient for span detection yet generally stop short of normalization, assertion/date attribution, or cross-sentence temporal reasoning, and their recall degrades as notes lengthen. By contrast, single-prompt LLM baselines mitigate some gaps (handling heterogeneous phrasing and implicit cues) but remain brittle for schema-aligned outputs: they do not reliably perform controlled-vocabulary normalization and are more prone to hallucination without systematic designs. In CLINES, a multi-step orchestration—chunking, tool-guided extraction, normalization, and date reconciliation—delivers structured outputs with transparent intermediate artifacts and maintains robustness across diverse note types. This positions CLINES as a scalable and sustainable approach for structuring clinical narratives. CLINES is positioned not as a competitor to contemporary production clinical-NLP suites or retrieval-augmented LLM pipelines but as a transparent, fully-specified, model-agnostic open reference implementation that prioritizes reproducibility of its prompt suite and an auditable intermediate representation. A fair matched comparison against those systems is left to future work; positioning details and related systems appear in Supplementary Materials.

### 4.4. Case studies: robust clinical inference

De-identified 4CE and CORAL notes show four recurring capabilities: inferring units when measurements are underspecified; normalizing terse or site-specific shorthand to controlled vocabularies; recovering relations that mirror care processes; and handling nuanced assertion status and qualitative values. Examples include resolving “Lat 2.4,” “BE –30,” and “Glic 617” to lactate, base excess, and glucose with appropriate units and UMLS links; mapping abbreviations such as IOT, TI, RX, PCI, and EVAR to procedures or imaging; linking coronary artery disease to s/p PCI; chaining biliary obstruction → stent placement → infectious management in oncology follow-ups; and retaining Present / Absent / Possible / Conditional assertions and qualitative ranges (e.g., “mid-90s” oxygen saturation) in machine-actionable form. These vignettes show the system operating as a clinical reasoner rather than a string matcher, reducing manual normalization burden while increasing fidelity for phenotyping and decision support in heterogeneous, length-variable notes.

### 4.5. Dataset-level variation and generalization

Performance varies between datasets for a given model and task, reflecting different writing styles and note sizes across disease areas. The multi-step, stepwise design limits error propagation: each module is independently validated, enabling interpretable error analysis. Semantic chunking is crucial for fitting long notes into LLM context windows, allowing even 4k–8k-token models to operate effectively. UMLS term normalization addresses LLM imprecision with controlled vocabularies, unifying semantically equivalent phrases (e.g., “heart attack” and “myocardial infarction”) under consistent codes and enhancing interoperability.

### 4.6. Schema adaptability

CLINES adapts output to target schemas (e.g., i2b2 star schema, registries, decision-support formats) and exports JSON, CSV, or SQLite, supporting integration with institutional analytics pipelines and EHR-linked databases.

### 4.7. Implications

For cohort discovery, phenotyping, and real-world evidence, CLINES can substantially reduce—though not eliminate—manual chart review for entities, assertions, values/units, and dates while leaving a verifiable audit trail. Given the residual hallucination rate quantified above, we recommend that adopters retain human verification for any extraction destined for safety-critical decisions. Sites can tailor prompts, dictionaries, and export schemas to local workflows without retraining large models.

### 4.8. Deployment, interoperability, and ethical considerations

Realistic deployment depends on factors outside the extraction algorithm itself: input preparation (PHI de-identification, segmentation), compute (FP8 8×H100 for Llama-3.1-405B; hosted-API for o3-mini/GPT-4o), and governance (institution-managed Azure deployments such as HMS Azure AI are scoped to de-identified data, and institutions handling identifiable PHI or GDPR-regulated data must select a corresponding HIPAA-compliant or in-region cloud configuration). UMLS CUIs are an ontology-grounded intermediate; downstream FHIR/OMOP-CDM use requires mapping to SNOMED CT, LOINC, or RxNorm. Demographic-attribute extraction is provided for cohort discovery and health-equity auditing only, and requires subgroup-stratified pre-deployment evaluation. Full discussion is provided in Supplementary Materials.

### 4.9. Limitations

Several limitations remain. (i) Prompt engineering influences performance at module level; prompts were not dataset- or model-optimized. (ii) Comparisons among o3-mini/GPT-4o and Llama-3.1-405B are not apples-to-apples given architecture, data, and token-limit differences. (iii) Hallucination risk persists, especially for ambiguous references or rare abbreviations (e.g., “OD 2 gtt qd” misread as overdose rather than ophthalmic dosing). Our seven-class taxonomy showed that the majority of apparent false positives are annotation gaps rather than model fabrications, with each true-hallucination category below 17% of audited cases. (iv) Gold annotations came from different annotators across datasets, introducing variability. Our cross-annotation study (mention F1 = 0.82; assertion *κ* = 0.85) bounds the human-agreement variability within which model-versus-gold differences should be interpreted. Future work should quantify inter-annotator agreement and explore ensemble annotation strategies. Additional limitations relating to note-type coverage (discharge summaries, operative and radiology reports), the absence of gold-standard relation annotations, and the unoptimised single-prompt and chain-of-thought baselines are addressed in Supplementary Materials.

### 4.10. Future work

Future directions include domain-adaptive fine-tuning (e.g., oncology, pediatrics), adaptive feedback loops where clinician corrections refine prompts or parameters, and multilingual extensions to support non-English healthcare systems. Other directions include hybrid per-stage model routing, retrieval-grounded extraction with span-level provenance, subgroup-stratified fairness evaluation, expansion to additional note types, and OMOP-CDM/FHIR integration; expanded discussion is in the Supplementary Materials.

### 4.11. Conclusion

CLINES translates clinical free text into ontology-grounded, attributed, time-stamped, and auditable structured data, with the gains over single-prompt LLMs and full-size clinical encoders attributable to the multi-step architecture rather than backbone choice. A residual hallucination rate quantified by stratified error analysis mandates human verification for safety-critical use; CLINES offers a model-agnostic, reproducible route to scale chart-review-like extraction for cohort discovery and real-world evidence.

## Contributors

Conceptualization & supervision: Tianxi Cai, Isaac Kohane. Methodology & software: Zongxin Yang, Hongyi Yuan, Raheel Sayeed, Amelia Li Min Tan (design); Zongxin Yang, Hongyi Yuan (implementation). Data curation & annotation: Enci Cai, Mohammed Moro, Xiudi Li, Zongxin Yang, Raheel Sayeed. Investigation & cross-annotation (for inter-annotator agreement): Enci Cai, Mohammed Moro. Formal analysis: Zongxin Yang, Hongyi Yuan, Huaiyuan Ying. Data access & verification: Zongxin Yang, Raheel Sayeed. The i2b2 web demonstrator: Nicholas Brown, Griffin Weber. Writing—original draft: Zongxin Yang, Raheel Sayeed. Writing—review & editing: all authors. Accountability: all authors approved the final manuscript, and accept responsibility for its accuracy and integrity. The corresponding author had final responsibility for the decision to submit.

## Data Sharing

Source clinical notes were accessed under data-use agreements and are not publicly shareable. The MIMIC-III dataset is available from PhysioNet^
2^ under the PhysioNet credentialed-access process, which requires completion of CITI *Human Subjects Research* training and signing of the dataset’s Data Use Agreement. The CORAL dataset is available from PhysioNet^3^ under the same credentialed-access framework, requiring completion of CITI *Data or Specimens Only Research* training and signing of the dataset’s Data Use Agreement, as specified by the dataset custodians. The 4CE clinical-notes subset used in this study is the de-identified “clinical notes, focusing on cases of patients with obesity and a history of COVID” collection hosted on the Harvard Medical School DBMI Data Portal;^4^ access is granted by the dataset custodians upon approval of the project-specific Data Use Agreement. General information about the 4CE consortium (Consortium for Clinical Characterization of COVID-19 by EHR) is available online.^5^ Cross-annotation files used for the inter-annotator agreement analysis (de-identified mention selections on six notes) are available from the corresponding author on reasonable request, subject to verification of the requestor’s credentials for the source dataset. No additional patient-level data were generated by this study. Study-derived prompt templates and a public inference reference implementation of CLINES (single-note inference with user-supplied Azure OpenAI credentials) are released under the MIT License on GitHub.^6^ An interactive, read-only i2b2 web demonstrator populated with CORAL results is also available online.^7^

## Ethics Approval

This study involved secondary analysis of fully de-identified electronic health record data. Because the analysis involves only fully de-identified secondary data that are not readily identifiable, it does not constitute human-subjects research under the U.S. Common Rule (45 CFR 46.104) and did not require Institutional Review Board approval or a formal exemption determination at the authors’ institution (Harvard Medical School); no patient contact, intervention, or access to identifiable private information occurred. The datasets were obtained and used under their respective data-use agreements: PhysioNet credentialed access for MIMIC-III (CITI Human Subjects Research training); PhysioNet credentialed access for CORAL (CITI Data or Specimens Only Research training); and a project-specific Data Use Agreement with the 4CE clinical-notes custodians at the Harvard Medical School DBMI Data Portal. All annotators completed the CITI training course required for each dataset and held the access credentials required for each dataset prior to data access. All large-language-model inference was performed via Harvard Medical School Information Technology’s managed Azure AI service (HMS Azure AI),^8^ an HMS-administered deployment providing project-scoped access to Microsoft Azure OpenAI models for Harvard data-security Level 4 projects; only de-identified clinical text (containing no HIPAA-regulated identifiers) was processed, and no text was retained or used to train provider models. Ethical approvals and data-use agreements for the original data collection were obtained by the respective data custodians.

## Data Availability

All data produced are available online at https://ai.nejm.org/doi/full/10.1056/AIdbp2300110,https://covidclinical.net, and https://physionet.org/content/mimiciii/1.4/.

## Footnotes

2 https://physionet.org/content/mimiciii/

3 https://physionet.org/content/curated-oncology-reports/1.0/

4 https://portal.dbmi.hms.harvard.edu/projects/4ce-data-set/

5 https://covidclinical.net/

6 https://github.com/celehs/clines

7 celehs.hms.harvard.edu/i2b2_demo

8 https://it.hms.harvard.edu/service/azure-ai

## References

1 Tayefi M, Ngo P, Chomutare T, Dalianis H, Salvi E, Budrionis A, et al. Challenges and opportunities beyond structured data in analysis of electronic health records. WIREs Computational Statistics. 2021 Feb;13(6). Available from: 10.1002/wics.1549.

2 Sushil M, Kennedy VE, Mandair D, Miao BY, Zack T, Butte AJ. CORAL: Expert-Curated Oncology Reports to Advance Language Model Inference. NEJM AI. 2024 Mar;1(4). Available from: 10.1056/aidbp2300110.

3 Kohane IS, Aronow BJ, Avillach P, Beaulieu-Jones BK, Bellazzi R, Bradford RL, et al. What Every Reader Should Know About Studies Using Electronic Health Record Data but May Be Afraid to Ask. Journal of Medical Internet Research. 2021 Mar;23(3):e22219. Available from: 10.2196/22219.

4 Kim MK, Rouphael C, McMichael J, Welch N, Dasarathy S. Challenges in and Opportunities for Electronic Health Record-Based Data Analysis and Interpretation. Gut and Liver. 2023 Oct;18(2):201–208. Available from: 10.5009/gnl230272.

5 Savova GK, Masanz JJ, Ogren PV, Zheng J, Sohn S, Kipper-Schuler KC, et al. Mayo clinical Text Analysis and Knowledge Extraction System (cTAKES): architecture, component evaluation and applications. Journal of the American Medical Informatics Association. 2010 Sep;17(5):507–513. Available from: 10.1136/jamia.2009.001560.

6 Aronson AR. Effective mapping of biomedical text to the UMLS Metathesaurus: the MetaMap program. In: Proceedings of the AMIA Symposium; 2001. p. 17.

7 Friedman C, Hripcsak G, DuMouchel W, Johnson SB, Clayton PD. Natural language processing in an operational clinical information system. Natural Language Engineering. 1995 Mar;1(1):83–108. Available from: 10.1017/S1351324900000061.

8 Pathak J, Kho AN, Denny JC. Electronic health records-driven phenotyping: challenges, recent advances, and perspectives. Journal of the American Medical Informatics Association. 2013 Dec;20(e2):e206–e211. Available from: 10.1136/amiajnl-2013-002428.

9 Lample G, Ballesteros M, Subramanian S, Kawakami K, Dyer C. Neural Architectures for Named Entity Recognition. In: Proceedings of the 2016 Conference of the North American Chapter of the Association for Computational Linguistics: Human Language Technologies (NAACL-HLT); 2016. p. 260–70.

10 Lee J, Yoon W, Kim S, Kim D, Kim S, So CH, et al. BioBERT: a pre-trained biomedical language representation model for biomedical text mining. Bioinformatics. 2020;36(4):1234–40.

11 Alsentzer E, Murphy J, Boag W, Weng WH, Jindi D, Naumann T, et al. Publicly Available Clinical BERT Embeddings. In: Proceedings of the 2nd Clinical Natural Language Processing Workshop; 2019. p. 72–8.

12 Achiam J, Adler S, Agarwal S, Ahmad L, Akkaya I, Aleman FL, et al. GPT-4 Technical Report. arXiv preprint arXiv:230308774. 2023. Available from: https://arxiv.org/abs/2303.08774.

13 Singhal K, Azizi S, Tu T, Mahdavi SS, Wei J, Chung HW, et al. Large language models encode clinical knowledge. Nature. 2023 Jul;620(7972):172–180. Available from: 10.1038/s41586-023-06291-2.

14 Brown TB, Mann B, Ryder N, Subbiah M, Kaplan J, Dhariwal P, et al. Language Models are Few-Shot Learners. arXiv preprint arXiv:200514165. 2020. Available from: https://arxiv.org/abs/2005.14165.

15 Yang X, Chen A, PourNejatian N, Shin HC, Smith KE, Parisien C, et al. GatorTron: A Large Clinical Language Model to Unlock Patient Information from Unstructured Electronic Health Records. arXiv preprint arXiv:220303540. 2022. Available from: https://arxiv.org/abs/2203.03540.

16 Rohanian O, Nouriborji M, Kouchaki S, Clifton DA. On the effectiveness of compact biomedical transformers. Bioinformatics. 2023 Feb;39(3). Avail-able from: 10.1093/bioinformatics/btad103.

17 Rohanian O, Nouriborji M, Jauncey H, Kouchaki S, Nooralahzadeh F, Clifton L, et al. Lightweight transformers for clinical natural language processing. Natural Language Engineering. 2024 Jan;30(5):887–914. Available from: 10.1017/S1351324923000542.

18 Uzuner Ö, South BR, Shen S, DuVall SL. 2010 i2b2/VA challenge on concepts, assertions, and relations in clinical text. Journal of the American Medical Informatics Association. 2011;18(5):552–6.

19 Radford A, Narasimhan K, Salimans T, Sutskever I. Improving Language Understanding by Generative Pre-Training. OpenAI; 2018. Available from: https://cdn.openai.com/research-covers/language-unsupervised/language_understanding_paper.pdf.

20 Karabacak M, Margetis K. Embracing Large Language Models for Medical Applications: Opportunities and Challenges. Cureus. 2023 May. Available from: 10.7759/cureus.39305.

21 Rawte V, Sheth A, Das A. A Survey of Hallucination in Large Foundation Models. arXiv preprint arXiv:230905922. 2023. Available from: https://arxiv.org/abs/2309.05922.

22 Zhao WX, Zhou K, Li J, Tang T, Wang X, Hou Y, et al. A Survey of Large Language Models. arXiv preprint arXiv:230318223. 2023. Available from: https://arxiv.org/abs/2303.18223.

23 Grattafiori A, Dubey A, Jauhri A, Pandey A, Kadian A, Al-Dahle A, et al. The Llama 3 Herd of Models. arXiv preprint arXiv:240721783. 2024. Available from: https://arxiv.org/abs/2407.21783.

24 Brat GA, Weber GM, Gehlenborg N, Avillach P, Palmer NP, Chiovato L, et al. International electronic health record-derived COVID-19 clinical course profiles: the 4CE consortium. npj Digital Medicine. 2020 Aug;3(1). Available from: 10.1038/s41746-020-00308-0.

25 OpenAI. tiktoken; 2024. GitHub repository, release 0.6.0 (published 2024-02-09); https://github.com/openai/tiktoken.

26 Isaacus. semchunk; 2024. GitHub repository, version 2.0.0 (released 2024-06-19); https://github.com/isaacus-dev/semchunk.

27 Liu F, Shareghi E, Meng Z, Basaldella M, Collier N. Self-alignment Pre-training for Biomedical Entity Representations. In: Proceedings of the 2021Conference of the North American Chapter of the Association for Computational Linguistics: Human Language Technologies; 2021. p. 4228–38.

28 He T, Belouali A, Patricoski J, Lehmann H, Ball R, Anagnostou V, et al. Trends and opportunities in computable clinical phenotyping: A scoping review. Journal of Biomedical Informatics. 2023 Apr;140:104335. Available from: 10.1016/j.jbi.2023.104335.

29 Hou J, Zhao R, Gronsbell J, Lin Y, Bonzel CL, Zeng Q, et al. Generate Analysis-Ready Data for Real-world Evidence: Tutorial for Harnessing Electronic Health Records With Advanced Informatic Technologies. Journal of Medical Internet Research. 2023 May;25:e45662. Available from: 10.2196/45662.

30 ohnson AEW, Pollard TJ, Shen L, Lehman LwH, Feng M, Ghassemi M, et al. MIMIC-III, a freely accessible critical care database. Scientific Data. 2016 May;3(1). Available from: 10.1038/sdata.2016.35.

